# Co-occurrence of alcohol use and *Schistosoma mansoni* infection: prevalence, patterns, and risk factors in rural Uganda

**DOI:** 10.1101/2025.10.14.25337972

**Authors:** Bethany Lyne, Micheal E. Besong, Betty Nabatte, Benjamin Tinkitina, John Bosco Oryema, Narcis B. Kabatereine, Sarah Lewington, Goylette F. Chami

## Abstract

**Background:** Despite being known independently as important aetiological agents of liver disease, little is known about the prevalence, patterns, and risk factors associated with co-occurring alcohol use and *Schistosoma mansoni* infection in rural endemic settings.

**Methods:** A cross-sectional analysis was completed of 3198 individuals aged 10-90 years from 52 rural villages in Uganda. Alcohol use was assessed using the World Health Organisation STEPwise Approach to Surveillance survey, and *S. mansoni* infection was diagnosed using Kato-Katz microscopy. Logistic regressions with biomedical, demographic, socioeconomic, and spatial variables were run.

**Results:** The overall prevalence of *S. mansoni* infection was approximately 44% (1405/3198) and current alcohol use was reported by over 12% (392/3198) of participants. Among males aged ≥20 years, the prevalence of co-occurring current alcohol use and *S. mansoni* infection was 18.7% (148/791). Females were less likely to have *S. mansoni* infection (odds ratio [OR] 0.83, 95% CI 0.69 – 0.99) or report current alcohol use (OR 0.44, 95% CI 0.32 – 0.62). Smoking (OR 9.43, 95% CI 6.66 – 13.37) and participating in fishing activity (OR 3.50, 95% CI 2.57 – 4.76) were associated with current alcohol use. Age (OR 0.95, 95% CI 0.94 – 0.97), fishing activity (OR 1.82, 95% CI 1.16 – 2.84), and smoking (OR 5.25, 95% CI 3.49 – 7.90) were associated with co-occurring current alcohol use and *S. mansoni* infection.

**Conclusion:** Co-occurrence of alcohol use and schistosome infection is common. Future research should investigate multi-sectoral public health interventions that simultaneously address the risk factors associated with both alcohol use and *S. mansoni* infection.

**Key messages:**

- This study aims to characterise the co-occurrence of alcohol use and *S. mansoni* infection, and to identify shared risk factors for these hepatotoxic exposures in a rural Ugandan population.
- Alcohol use was most prevalent among males aged ≥20 years, and key risk factors for both alcohol use and *S. mansoni* infection were male sex, current smoking status, and participation in fishing activity.
- The findings of this study highlight the need for integrated public health interventions that simultaneously target alcohol use and *S. mansoni* infection, focusing on high-risk occupational groups such as those involved with fishing and high-risk behaviours such as smoking.

## INTRODUCTION

Schistosomiasis and alcohol use are both prevalent in rural African settings, representing a double burden of infectious diseases and risk factors for non-communicable disease (NCDs) [1].

Understanding the co-distribution of schistosomiasis and alcohol use, particularly heavy drinking and chronic *Schistosoma mansoni* infection, is important for understanding liver injury [2, 3].

Schistosomiasis and alcohol use have overlapping populations at-risk. Schistosomiasis is a neglected tropical disease caused by infection with *Schistosoma* blood fluke parasites, which are transmitted indirectly to humans through contact with contaminated water sources with suitable intermediate snail hosts [4]. Over 250 million people are estimated to be infected with *Schistosoma* species worldwide, with 90% of them living in sub-Saharan Africa (SSA) [5, 6]. *S. mansoni*, which causes intestinal schistosomiasis, is among the most prevalent species in SSA [4]. Untreated chronic infections may cause liver morbidity, most notably periportal fibrosis, which can ultimately be fatal [7]. Alcohol use is also common in most parts of SSA, including areas where schistosomiasis is endemic [8]. For example, Uganda, which has an estimated national prevalence of 25% for *S. mansoni* [9], has one of highest levels of alcohol consumption in East Africa, with an annual per capita alcohol consumption of 26 litres, along with a prevalence of current alcohol users of varying frequency and intensity of 31.1% [10]. In rural fishing communities with high rates of water contact in Uganda, the prevalence of current alcohol use has been reported to be as high as 45% [10]. Alcohol consumption results in alcoholic liver disease (ALD), which represents a spectrum of conditions, starting with reversible steatosis, and progressing to more severe manifestations such as cirrhosis and hepatocellular carcinoma [11].

Although schistosomiasis and alcohol use have been individually studied in SSA, their co-occurrence has been rarely described [10]. While *S. mansoni* infection peaks in young children, alcohol use in Uganda is most prevalent among individuals in early to mid-adulthood (18 – 39 years) [12]. Despite this age disparity, studies have independently suggested that both schistosomiasis and alcohol use share risk factors, including male sex and participation in fishing activity [13–15]. The intersection of these factors, and the potential implications of their co-occurrence in the same population, remain understudied. Broader risk factors, such as behavioural and lifestyle patterns, that may drive this co-occurrence remain largely underexplored.

We investigated the co-occurrence of alcohol use and *S. mansoni* infection among 3198 individuals aged 10 years and older from 52 villages in Eastern and Western Uganda. We examined factors associated with current alcohol use and current *S. mansoni* infection individually, and then for their co-occurrence in a subset of the study population focused on male adults. The aim of this study was to describe the patterns of co-occurrence and identify shared biomedical, sociodemographic, behavioural, household, and spatial factors associated with both outcomes.

## METHODS

### Outcomes

We conducted a cross-sectional analysis of all individuals from the SchistoTrack cohort aged one year and older at the year-of-enrolment. Cohort details are provided elsewhere [16]. The primary aim of this study was to examine the co-occurrence of alcohol use and *S. mansoni* infection, with the main outcome of interest being co-occurring current alcohol use and *S. mansoni* infection. Secondary outcomes included current alcohol use and *S. mansoni* infection individually, which were analysed to understand the context of the co-occurrence model. Regular alcohol use and co-occurring regular alcohol use and *S. mansoni* infection were also examined, due to the known association between higher levels of alcohol use and the exacerbation of liver pathology [17].

*S. mansoni* infection status and intensity were assessed for participants aged ≥5 years using methods described previously with single stool samples and two thick smear slides [18]. Infection status was binary and defined as one or more eggs per gram (EPG). For descriptive analyses only, infection intensity was categorised according to current WHO guidelines as uninfected (0 EPG), low (1-99 EPG), moderate (100-399 EPG), and high (≥400 EPG) [19].

Alcohol use was assessed for participants aged ≥10 years using a modified version of the World Health Organisation STEPwise Approach to Surveillance (WHO STEPS) survey [20], administered as part of the household survey. Individuals identified as intoxicated, or incapacitated due to alcohol at the time of the household survey, were excluded from the study. Current alcohol use was a binary variable, coded as one if the participant reported alcohol use in the past 12 months. Regular alcohol use, also binary, was defined as alcohol consumption on a weekly or daily basis. Additional details on the collection of alcohol data are provided in the Supplementary Methods.

Co-occurring current alcohol use and *S. mansoni* infection was coded as a binary variable where an individual was positive if they were both categorised as a current alcohol user and had positive infection status. A similar binary variable was created for the co-occurrence of regular alcohol use and *S. mansoni* infection status. Co-occurring heavy *S. mansoni* infection and current alcohol use, and heavy *S. mansoni* infection and regular alcohol use are presented descriptively only rather than analysed as secondary model outcomes due to limited observations.

### Covariates

This study analysed variables from SchistoTrack that have been identified in the literature as potential risk factors or confounders for alcohol use or *S. mansoni* infection. Biomedical factors were self-reported praziquantel use, human immunodeficiency virus (HIV) infection, and history of depression (all binary). Sociodemographic factors comprised age in years (count), sex (binary), belonging to the majority religion of the village (binary), belonging to the majority tribe of the village (binary), years of educational attainment (count), and occupation (categorical, with options such as fishing, fishmongering, subsistence farming and rice farming that constituted the main income-earning occupation of the individual). The majority-religion and majority-tribe variables were constructed instead of specific religious or tribal identifiers to better capture the impacts of social exclusion and to avoid stigmatisation of a particular group. The broader district variable was expected to capture differences related to culture or religion in the community. Behavioural factors were fishing activity, fishmongering activity, and current smoking within the past 12 months (all binary). Fishing and fishmongering activity variables were assessed based on weekly engagement with water sources, regardless of them constituting a formal income-earning occupation (hence including also children), and so were included in this analysis instead of occupation. Household-level variables included home quality score (continuous), along with water and sanitation indicators: improved water source, water purification, improved sanitation, and hygiene facility (all binary). All household variables have been defined elsewhere [21, 22], apart from hygiene facility, which was considered present if soap or detergents were available at handwashing locations.

### Statistical analysis

To improve the generalisability of our results, direct age standardisation was carried out using the Ugandan National Housing and Population Census 2024 as the standard population [23]. Age standardisation was carried out in Microsoft Excel (v2108).

All models were run using RStudio (version 4.2). Multivariable logistic regressions were performed to analyse the associations between covariates and *S. mansoni* infection, current alcohol use, and regular alcohol use. No variable selection was performed on these outcomes. All participants were included in these analyses, with robust standard errors clustered by household to account for paired sampling of individuals within a household [24]. Due to the majority of alcohol use being observed in males aged ≥20 years, analyses of co-occurrence were restricted to this subgroup. For the subgroup analysis, Bayesian variable selection was performed using the Bayesian Adaptive Sampling (BAS) package in R, where a uniform prior distribution was applied, assigning equal probability to all possible models [25]. All covariates with inclusion probabilities greater than 0.5 were used to build the subgroup models. For categorical variables with more than two categories, floating absolute risks (FAR) were calculated to allow confidence intervals (CIs) to be estimated for all groups, and reported to assess trends in heterogeneity [26]. This method, based on the variance of the log odds, avoids dependence on an arbitrarily chosen reference category and improves interpretability [27]. Variance inflation factors (VIFs) were used to test for multicollinearity between variables [28]. A VIF level > 5 was considered suggestive of multicollinearity; variables with a VIF greater than this would be removed from the model and run separately. For all regression models, the optimum probability cut-off threshold for classification was determined by maximising Youden’s index (J), using the pROC package in R, based receiver operating characteristic (ROC) curve and its area under the curve (auROC) [29]. To assess predictive capacity for all models, 10-fold cross validation (5-fold for the subgroup analyses) was used [30].

## RESULTS

### Participant characteristics

After exclusions (Figure 1), 3198 individuals had data at enrolment on both *S. mansoni* infection and alcohol use, of which 24.73% (791) were males aged ≥20 years. The overall prevalence of *S. mansoni* infection was 43.9% (1405/3198). Current alcohol use was reported by 12.3% (392/3198) of participants, and regular alcohol use by 6.7% (214/3198) (Table 1). Age standardised prevalence rates were 45.5% for schistosome infection, 11.7% for current alcohol use, and 6.4% for regular alcohol use (Supplementary Figures S1-S2). Among those who consumed alcohol, 52.8% (207/392) reported drinking homebrewed alcohol, 22.2% (87/392) beer, and 11.7% (46/392) spirits. The median age of participants was 30 (IQR 14 – 43) years, with an age range of 10 – 90 years, reflecting the eligibility criterion of ≥10 years for alcohol use data collection. Among the 3198 participants, 56.8% (1818/3198) were female, 17.5% (560/3198) reported participating in fishing activity, and 8.3% (266/3198) reported current smoking. The most frequently reported religion was Christianity (74.4%, 2376/3198), and the three major tribes were Alur (61.8%, 1972/3198), Musoga (12.2%, 391/3198), and Bagungu (8.9%, 285/3198). Tribe and district showed collinearity – 65.1% (1284/1972) of Alur participants resided in Pakwach District, 100% (391/391) of Musoga participants resided in Mayuge district, and 90.2% (257/285) of Bagungu participants resided in Buliisa district.

**Figure 1.**
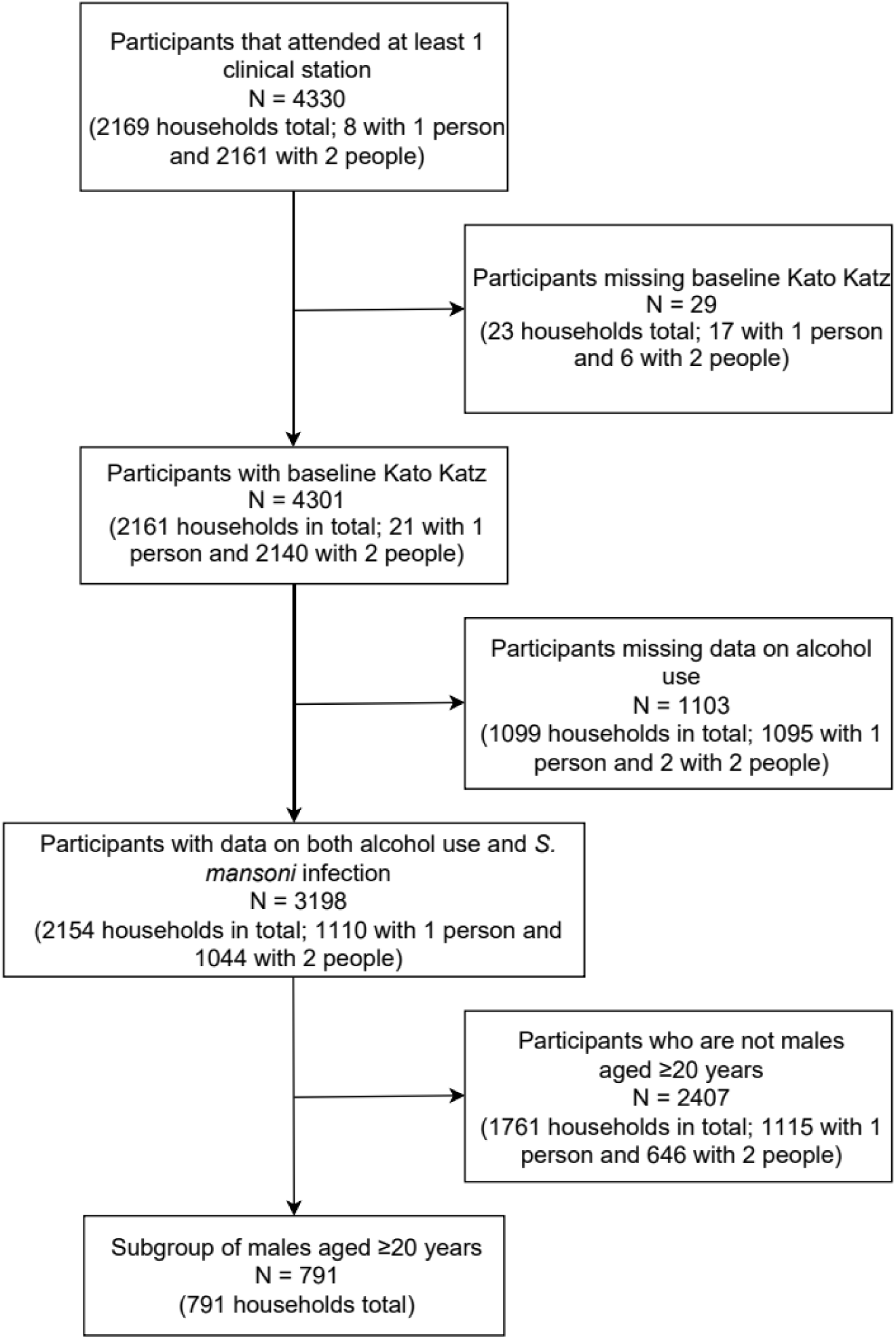
Participant flowchart

**Table 1.** Baseline characteristics of the SchistoTrack cohort, overall and by data availability.

| Characteristic | All participants<br>(N = 8042) | Alcohol data<br>(N = 5857) | Infection data<br>(N = 4301) | Alcohol and<br>infection data<br>(N = 3198) | Males aged 20+<br>(N = 791) |
| --- | --- | --- | --- | --- | --- |
| **OUTCOMES** |  |  |  |  |  |
| Alcohol use (n, %) |  |  |  |  |  |
| Alcohol consumption (ever) | 707 (8.8) | 707 (12.1) | 439 (10.2) | 439 (13.7) | 317 (40.0) |
| Former alcohol user | 73 (0.9) | 73 (1.2) | 47 (1.1) | 47 (1.5) | 22 (2.8) |
| Occasional alcohol user | 288 (3.6) | 288 (4.9) | 178 (4.1) | 178 (5.6) | 118 (14.9) |
| Regular alcohol user | 346 (4.3) | 346 (5.9) | 214 (5.0) | 214 (6.7) | 177 (22.4) |
| *S. mansoni* infection (n, %) | 1785 (22.2) | 1405 (24.0) | 1785 (41.5) | 1405 (43.9) | 356 (45.0) |
| **BIOMEDICAL FACTORS** |  |  |  |  |  |
| Praziquantel intake (n, %) | 3550 (44.1) | 2975 (50.8) | 1968 (45.8) | 1645 (51.4) | 447 (56.5) |
| HIV (n, %) | 127 (1.6) | 114 (1.9) | 78 (1.8) | 70 (2.2) | 19 (2.4) |
| Depression (n, %) | 102 (1.3) | 92 (1.6) | 70 (1.6) | 65 (2.0) | 17 (2.1) |
| **SOCIODEMOGRAPHIC FACTORS** |  |  |  |  |  |
| Age |  |  |  |  |  |
| Median (IQR)* | 17.0 (9.0 – 35.0) | 28.0 (15.0 – 40.0) | 18.0 (9.0 – 38.0) | 30.0 (14.0 – 43.0) | 39.0 (31.0 – 49.0) |
| Sex (n, %) |  |  |  |  |  |
| Male | 3816 (47.5) | 2698 (46.1) | 1962 (45.7) | 1380 (43.2) | 791 (100) |
| Female | 4226 (52.5) | 3159 (53.9) | 2339 (54.3) | 1818 (56.8) |  |
| Religion (n, %) |  |  |  |  |  |
| Christian | 5862 (72.8) | 4307 (73.6) | 3178 (73.9) | 2376 (74.4) | 589 (74.5) |
| Born-again Christian | 964 (12.0) | 695 (11.9) | 519 (12.1) | 380 (11.9) | 81 (10.2) |
| Muslim | 1122 (13.9) | 780 (13.3) | 552 (12.8) | 400 (12.5) | 106 (13.4) |
| No religion | 19 (0.2) | 15 (0.3) | 11 (0.3) | 9 (0.3) | 4 (0.5) |
| Other | 75 (0.9) | 60 (1.0) | 41 (1.0) | 33 (1.0) | 11 (1.4) |
| Tribe (n, %) |  |  |  |  |  |
| Alur | 4739 (58.9) | 3516 (60.0) | 2628 (61.1) | 1972 (61.8) | 523 (66.1) |
| Musoga | 1081 (13.4) | 759 (12.9) | 527 (12.3) | 391 (12.2) | 70 (8.9) |
| Bagungu | 667 (8.3) | 478 (8.2) | 408 (9.5) | 285 (8.9) | 93 (11.7) |
| Others | 1555 (19.3) | 1104 (18.9) | 738 (17.2) | 550 (17.2) | 105 (13.3) |
| Highest level of education attained |  |  |  |  |  |
| Median (IQR)* | 3.0 (1.0 – 6.0) | 4.0 (2.0 – 6.0) | 3.0 (1.0 – 5.0) | 4.0 (2.0 – 6.0) | 6.0 (4.0 – 7.0) |
| Occupation (n, %) |  |  |  |  |  |
| None | 2813 (35.0) | 1585 (27.1) | 1388 (32.3) | 788 (24.6) | 103 (13.0) |
| Housewife | 454 (5.6) | 412 (7.0) | 212 (4.9) | 209 (6.5) | 4 (0.5) |
| Student | 1763 (21.9) | 1045 (17.8) | 1063 (24.8) | 588 (18.4) | 6 (0.8) |
| Rice farmer | 20 (0.2) | 20 (0.3) | 11 (0.3) | 11 (0.3) | 5 (0.6) |
| Subsistence farmer | 1157 (14.4) | 1140 (19.5) | 677 (15.7) | 668 (20.9) | 163 (20.6) |
| Fisherman | 611 (7.6) | 608 (10.4) | 311 (7.2) | 309 (9.7) | 299 (37.8) |
| Fishmonger | 220 (2.7) | 220 (3.8) | 145 (3.4) | 145 (4.5) | 18 (2.3) |
| Other | 1004 (12.5) | 827 (14.1) | 494 (11.5) | 480 (15.0) | 193 (24.4) |
| Home quality score |  |  |  |  |  |
| Median (IQR)* | 3.0 (3.0 – 9.0) | 3.0 (3.0 – 9.0) | 3.0 (3.0 – 9.0) | 3.0 (3.0 – 9.0) | 3.0 (3.0 – 8.5) |
| **BEHAVIOURAL FACTORS** |  |  |  |  |  |
| Fishing activity (n, %) | 1025 (12.7) | 1005 (17.2) | 571 (13.3) | 560 (17.5) | 463 (58.5) |
| Fishmonger activity (n, %) | 282 (3.5) | 277 (4.7) | 197 (4.6) | 193 (6.0) | 14 (1.8) |
| Current smoker (n, %) | 454 (5.6) | 453 (7.7) | 267 (6.2) | 266 (8.3) | 204 (25.8) |
| **HOUSEHOLD WASH FACTORS** |  |  |  |  |  |
| Improved water source (n, %) | 4260 (53.0) | 3082 (52.6) | 2229 (51.9) | 1687 (52.7) | 418 (52.8) |
| Water purification (n, %) | 1792 (22.3) | 1341 (22.9) | 963 (22.4) | 734 (22.9) | 200 (25.3) |
| Improved sanitation (n, %) | 5821 (72.3) | 4247 (72.4) | 3099 (72.0) | 2302 (72.0) | 594 (75.1) |
| Basic hygiene facility (n, %) | 799 (9.9) | 590 (10.1) | 406 (9.5) | 305 (9.5) | 96 (12.1) |
| **SPATIAL FACTORS** |  |  |  |  |  |
| District (n, %) |  |  |  |  |  |
| Buliisa | 2456 (30.6) | 1766 (30.2) | 1458 (33.9) | 1030 (32.2) | 248 (31.4) |
| Mayuge | 2270 (28.2) | 1590 (27.1) | 1074 (25.0) | 795 (24.9) | 148 (18.7) |
| Pakwach | 3316 (41.2) | 2501 (42.7) | 1769 (41.1) | 1373 (42.9) | 395 (49.9) |

Acronyms: IQR – interquartile range; HIV – human immunodeficiency virus; WASH – water sanitation and hygiene.

Table 2 shows the prevalence of each outcome stratified by age, sex, and district. Overall, the prevalence of co-occurring current alcohol use and *S. mansoni* infection was 5.7% (183/3198), while co-occurring regular alcohol use and *S. mansoni* infection was observed in 3.4% (108/3198) of participants. Among males aged ≥20 years, the prevalence of co-occurring current alcohol use and *S. mansoni* infection was 18.7% (148/791), and for co-occurring regular alcohol use and *S. mansoni* infection the prevalence was 11.3% (89/791). Overall, males aged ≥20 years accounted for 75.3% (295/392) of current alcohol use.

**Table 2.** Prevalence of alcohol use, *S. mansoni* infection, and their co-occurrence, stratified by age, sex, and district.

|  |  | Alcohol Use |  |  |  |  |  |  |
| --- | --- | --- | --- | --- | --- | --- | --- | --- |
|  |  | Current alcohol non-users |  | Current alcohol users |  |  |  |  |
|  | N | Lifetime alcohol non-users (n, %) | Former alcohol users (n, %) | Occasional alcohol users (n, %) | Regular alcohol users (n, %) | *S. mansoni* infection at baseline (n, %) | Current alcohol use/*S. mansoni* co-occurrence (n, %) | Regular alcohol use/*S. mansoni* co-occurrence (n, %) |
| Overall | 3198 | 2759 (86.3) | 47 (1.5) | 178 (5.6) | 214 (6.7) | 1405 (43.9) | 183 (5.7) | 108 (3.4) |
| Males |  |  |  |  |  |  |  |  |
| Total | 1380 | 1052 (76.3) | 24 (1.7) | 125 (9.1) | 179 (13.0) | 693 (50.2) | 150 (10.9) | 90 (6.5) |
| Age (years) |  |  |  |  |  |  |  |  |
| 10 – 19 | 589 | 578 (98.1) | 2 (0.3) | 7 (1.2) | 2 (0.3) | 337 (57.3) | 2 (0.3) | 1 (0.2) |
| 20 – 29 | 144 | 79 (54.9) | 3 (2.1) | 23 (16.0) | 39 (27.1) | 95 (65.9) | 43 (29.9) | 26 (18.1) |
| 30 – 39 | 257 | 156 (60.7) | 4 (1.6) | 40 (15.6) | 57 (22.2) | 131 (51.0) | 51 (19.9) | 31 (12.1) |
| ≥40 | 390 | 239 (61.3) | 15 (3.8) | 55 (14.1) | 81 (20.8) | 130 (33.3) | 54 (13.8) | 32 (8.2) |
| District |  |  |  |  |  |  |  |  |
| Mayuge | 282 | 256 (90.8) | 4 (1.4) | 19 (6.7) | 3 (1.1) | 135 (47.9) | 12 (4.3) | 3 (1.1) |
| Buliisa | 408 | 307 (75.3) | 4 (1.0) | 34 (8.3) | 63 (15.4) | 179 (43.9) | 42 (10.3) | 27 (6.6) |
| Pakwach | 690 | 489 (70.9) | 16 (2.3) | 72 (10.4) | 113 (16.3) | 379 (54.9) | 96 (13.9) | 60 (8.7) |
| Females |  |  |  |  |  |  |  |  |
| Total | 1818 | 1707 (93.9) | 23 (1.3) | 53 (2.9) | 35 (1.9) | 712 (39.2) | 33 (1.8) | 18 (1.0) |
| Age (years) |  |  |  |  |  |  |  |  |
| 10 – 19 | 486 | 482 (99.2) | 1 (0.2) | 1 (0.2) | 2 (0.4) | 256 (52.7) | 1 (0.2) | 1 (0.2) |
| 20 – 29 | 340 | 327 (96.2) | 1 (0.3) | 7 (2.1) | 5 (1.5) | 141 (41.5) | 7 (2.1) | 3 (0.9) |
| 30 – 39 | 392 | 359 (91.6) | 6 (1.5) | 16 (4.1) | 11 (2.8) | 151 (38.5) | 12 (3.1) | 8 (2.0) |
| ≥40 | 600 | 539 (89.8) | 15 (2.5) | 29 (4.8) | 17 (2.8) | 164 (27.3) | 13 (2.2) | 6 (1.0) |
| District |  |  |  |  |  |  |  |  |
| Mayuge | 513 | 489 (95.3) | 2 (0.4) | 20 (3.9) | 2 (0.4) | 152 (29.6) | 6 (1.2) | 0 (0.0) |
| Buliisa | 622 | 583 (93.7) | 5 (0.8) | 11 (1.8) | 23 (3.7) | 273 (43.9) | 20 (3.2) | 14 (2.2) |
| Pakwach | 683 | 635 (93.0) | 16 (2.3) | 22 (3.2) | 10 (1.5) | 287 (42.0) | 7 (1.0) | 4 (0.6) |

### Covariates associated with *S. mansoni* infection

Figure 2 shows the results of the logistic regression models for *S. mansoni* infection. In the fully adjusted model, female participants had 0.83 times the odds of infection compared to males (95% CI 0.69 – 0.99). Strong associations were observed for behavioural exposures. Participants who engaged in fishing activity had 1.44 times the odds of infection compared to those who did not (95% CI 1.14 – 1.81), and current smokers had 1.58 times the odds of infection compared to current non-smokers (95% CI 1.17 – 2.11). Based on the FAR, district-level differences were observed. Residents of Pakwach district had 1.62 times the odds of infection, and residents of Buliisa district had 1.22 times the odds (95% CI 1.09 – 1.37) compared to the overall average. The lack of overlap in the 95% CIs for Pakwach indicates that the odds of infection in this district are significantly higher than in the other districts.

**Figure 2.**
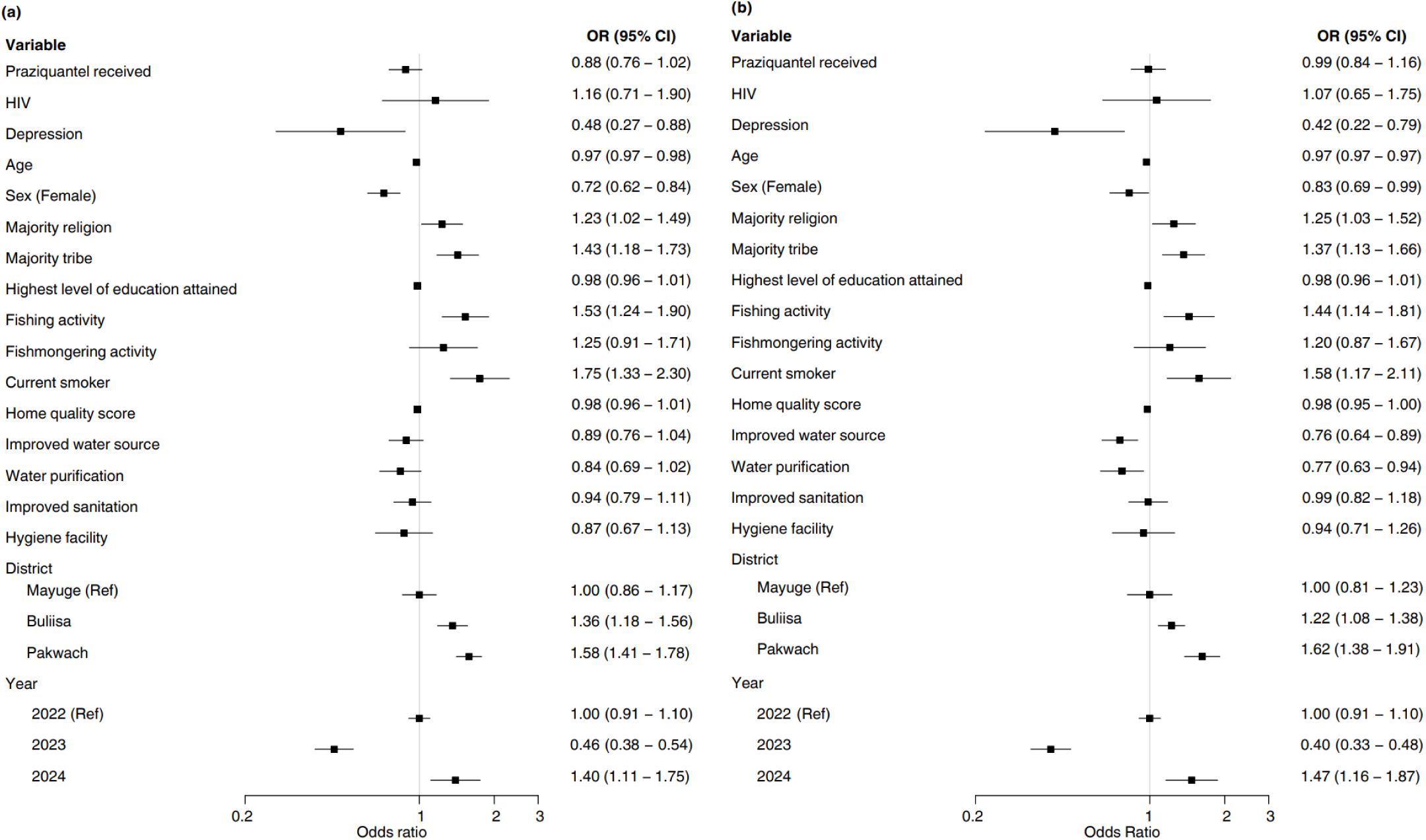
Models for *S. mansoni* infection status. Logistic regression models for *S. mansoni* infection status (1405/3198), with floating absolute risks (FAR) shown for district and year variables. (a) Minimally adjusted model, with each variable adjusted for age, sex, and district as appropriate. (b) Fully adjusted model including all covariates. Variance inflation factors (VIFs) were <5 for all variables. AUC for 10-fold cross-validation of the fully adjusted model was 0.681.

### Covariates associated with current alcohol use

Figure 3 shows the results of the logistic regression models for current alcohol use. Similar to *S. mansoni* infection status, females had lower odds of reporting current alcohol use compared to males (odds ratio [OR] 0.44, 95% CI 0.32 – 0.62). Current smokers had 9.43 times the odds of reporting current alcohol use compared to current non-smokers (95% CI 6.66 – 13.37). Participants who participated in fishing activity had 3.50 times higher odds of reporting current alcohol use compared to those who did not (95% CI 2.57 – 4.76). The geographical associations with district in the minimally adjusted model were attenuated and no longer significant in the fully adjusted model, as shown by the overlapping CIs of the FAR. The model had high predictive accuracy (mean auROC 0.87) Similar results were obtained in the logistic regression models for regular drinking (Supplementary Figure S3).

**Figure 3.**
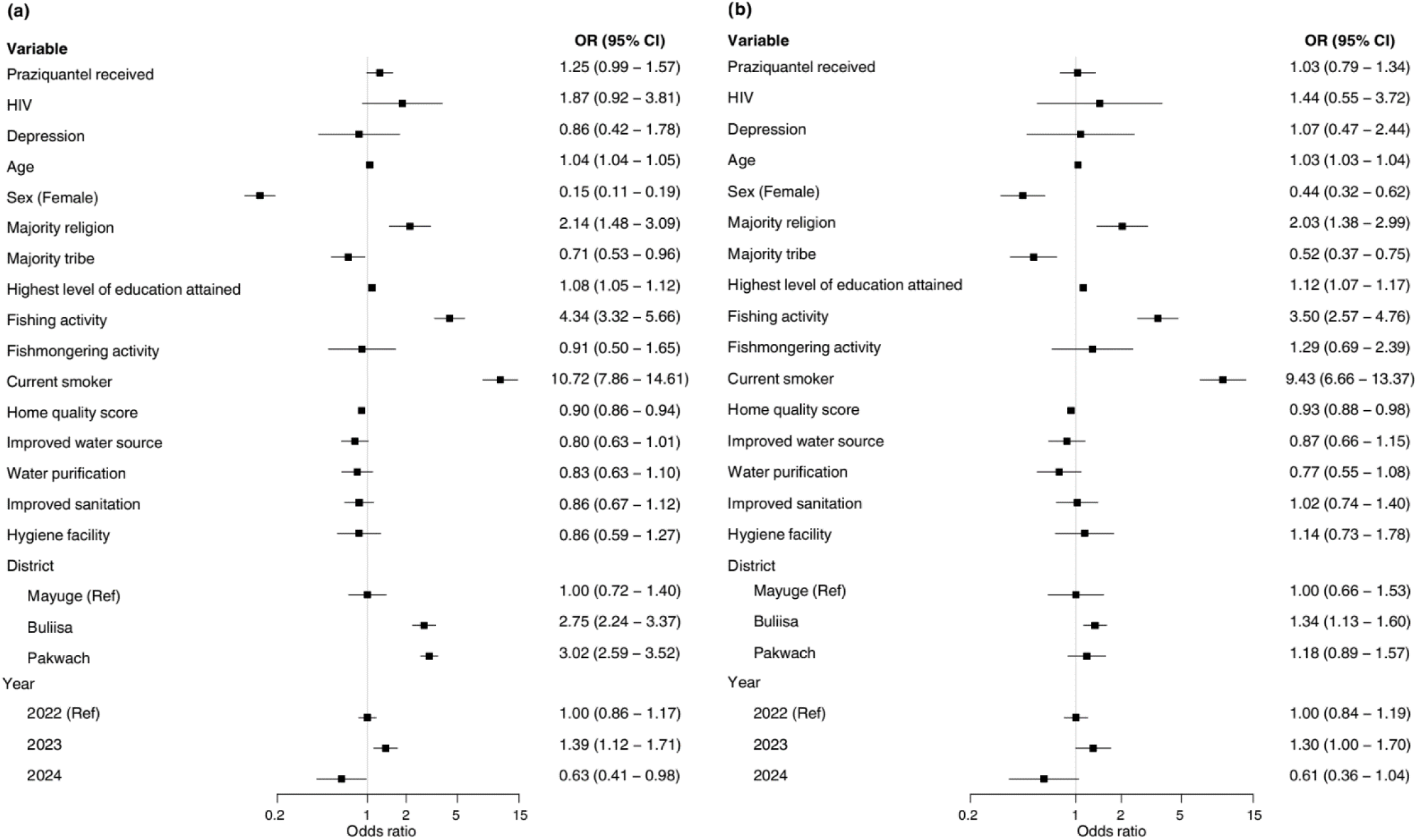
Models for current alcohol use. Logistic regression models for current alcohol use (392/3198), with floating absolute risks (FAR) shown for the district and year variables. (a) Minimally adjusted model, with each variable adjusted for age, sex, and district as appropriate. (b) Fully adjusted model including all covariates. Variance inflation factors (VIFs) were <5 for all variables. AUC for 10-fold cross-validation of the fully adjusted model was 0.868.

### Covariates associated with current drinking and *S. mansoni* co-occurrence

For males aged ≥20 years, only three variables were selected in the model for co-occurring current alcohol use and *S. mansoni* infection status: age, fishing activity and current smoking. The model demonstrated high predictive accuracy (auROC 0.77, Figure 4), with inclusion probabilities ranging from 0.65 to 0.99. A five-year increase in age was associated with 0.77 times lower odds of co-occurring current alcohol use and *S. mansoni* infection (95% CI 0.73 – 0.86). Participation in fishing activity was associated with 1.82 times higher odds compared to those who did not (95% CI 1.16 – 2.84), while current smokers had 5.25 times higher odds (95% CI 3.49 – 7.90) compared to non-smokers. Similar results were observed for co-occurring regular alcohol use and *S. mansoni* infection status, except for improved water source selected instead of fishing activity (Supplementary Figure S4). The distribution of *S. mansoni* infection intensity across the alcohol consumption groups among males aged ≥20 years is presented in Supplementary Table S5. A significant but weak association was found between infection intensity and likelihood of current alcohol use (χ^2^ 9.15, p = 0.17; Spearman’s ρ = 0.093, p-value = 0.0091).

**Figure 4.**
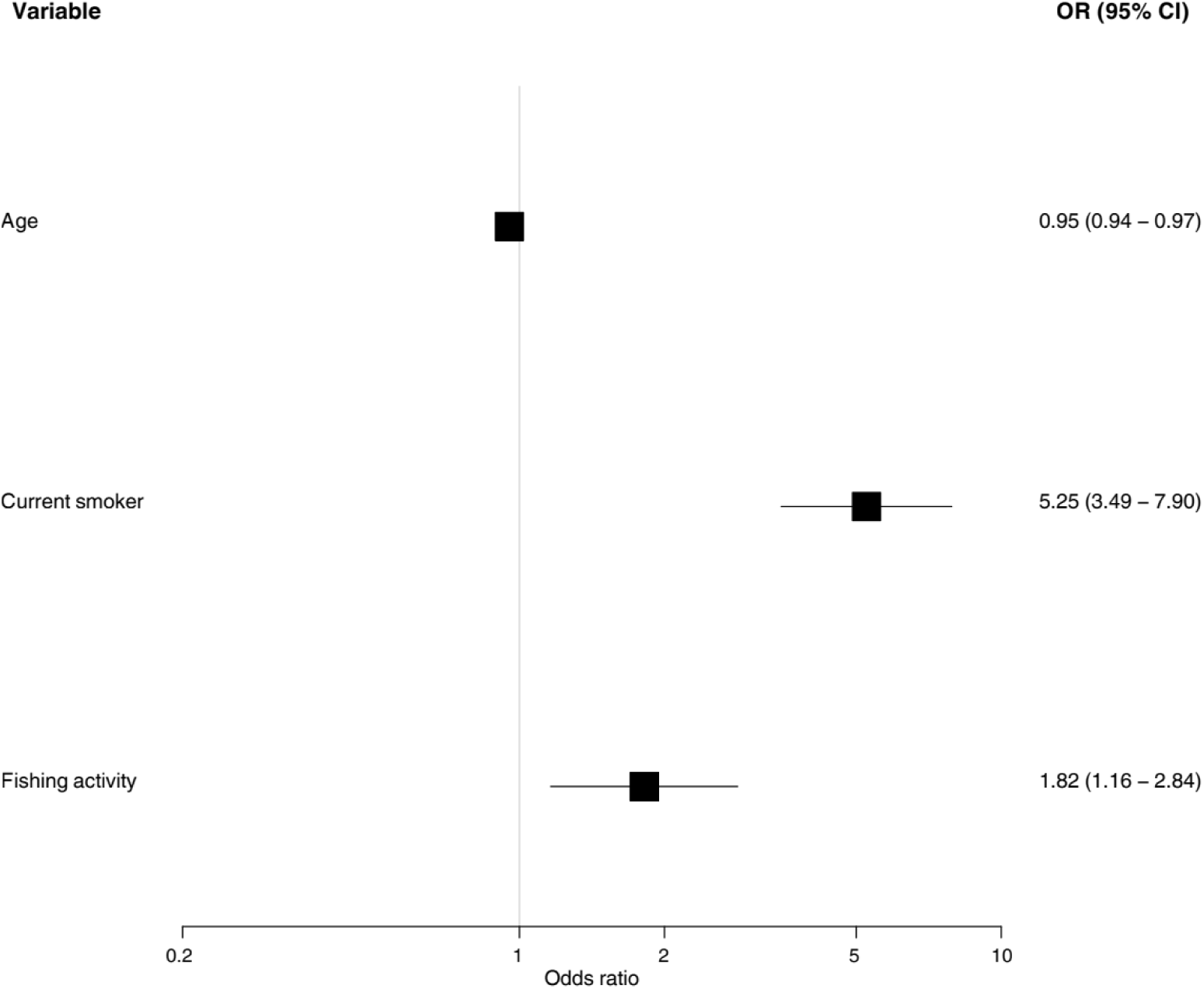
Model for co-occurring current alcohol use and *S. mansoni* infection. Logistic regression model for co-occurring current alcohol use and *S. mansoni* infection among males aged ≥20 years (148/791). VIFs<5 for all variables. AUC for 5-fold cross-validation was 0.774.

## DISCUSSION

Given the growing double burden of infectious and NCDs in low- and middle-income countries [31], this study aimed to identify the shared risk factors for the co-occurrence of a major risk factor for NCDs (current alcohol use) and *S. mansoni* infection status. We conducted a large-scale study in rural Uganda within 52 villages in the SchistoTrack cohort, analysing 3198 individuals aged ≥10 years. Overall, *S. mansoni* infection was identified in 43.9% of participants, 12.3% of participants reported current alcohol use, and 18.7% of males aged ≥20 years had co-occurring current alcohol use and *S. mansoni* infection. Drinking behaviours were most common in men, and over half of males who reported current alcohol use were regular alcohol users. Demographic, lifestyle, and occupational factors were discovered to be shared predictors of current alcohol use and *S. mansoni* infection status.

Male sex was a consistent shared factor associated with both *S. mansoni* infection and alcohol use, and can be linked to several cultural and occupational factors. Socially, alcohol consumption for men is viewed as a sign of independence and a way to bond with peers [32]. Many men drink in public spaces such as bars, where it is treated as a communal activity. In contrast, alcohol use among women is stigmatised due to the perception that it signifies neglect of domestic responsibilities and challenges gender norms [32]. This social context is compounded by occupational factors, as fishing is a male-dominated occupation in this setting. The necessity of frequent water contact increases the risk of *S. mansoni* infection, and the nature of fishing work – characterised by physical danger, psychological stress, and an irregular cash income – induces alcohol use as a way of coping [14,33]. Local infrastructure in Uganda also facilitates alcohol use; it was reported in a qualitative study that bars are frequently located near fishing sites and have long opening hours, which normalises alcohol use [14]. Given the overlapping locations of increased alcohol use and schistosome infection risk, future health education or outreach campaigns should consider designing interventions around fishing sites. Importantly, the identification of fishermen (and adult males generally) as a high-risk group provides a clear target for clinical research, particularly to assess the contributions of alcohol use and *S. mansoni* infection to liver pathologies including fatty liver disease and fibrosis.

Smoking status, another significant predictor, has consistently been linked to alcohol consumption, as epidemiological studies often report clustering of these behaviours [34, 35]. This is due to both behaviours being driven by similar social and psychological motivations, such as stress [36]. Additionally, access to improved water sources, which also reflects broad preventative health mechanisms, was associated with reduced odds of co-occurring regular alcohol use and *S. mansoni* infection, further showing the clustering of health-related behaviours. This analysis also revealed geographical patterns for different levels of alcohol use. For current alcohol use, no significant district-level associations were observed. However, in the regular alcohol use model, dramatically elevated odds were observed for Pakwach and Buliisa districts in the minimally adjusted model. These associations were substantially attenuated after adjustment for fishing activity and current smoking status, indicating significant confounding by these factors. This pattern suggests that while geography is not a determinant of any alcohol use, it may be associated with regular alcohol use through differences in infrastructure, cultural and religious practices, and the concentration of fishing activity.

While the overall prevalence rates of current and regular alcohol use observed in this study are lower than those reported in previous studies in rural African settings and nationwide surveys in Uganda [10, 37], this is likely due to the inclusion of participants as young as 10 years old, who diluted the overall prevalence estimates. When considering the demographic of males aged ≥20 years, the prevalence rates increased to 37.3% for current alcohol use and 22.4% for regular alcohol use, figures which are more comparable to those reported in previous studies of similar high-risk groups in Uganda [10]. This shows how the burden of alcohol use in these communities is concentrated among adult men, a factor which should be considered when implementing public health interventions.

This study has several strengths. We analysed a large community-based cohort with representativeness of schistosomiasis-endemic areas, a wide age range, and an ethnically and socially diverse population. We provide statistically robust models with a comprehensive set of covariates adjusted, each with good predictive capacity. However, the study has some limitations. Social desirability bias may have influenced participant responses, resulting in an underestimation of the true prevalence of alcohol consumption. Additionally, the method of data collection, where the household head reports alcohol consumption on behalf of other members, may have caused mis-reporting of alcohol use. Moreover, SchistoTrack excludes individuals who were heavily intoxicated and unable to perform daily tasks at the time of recruitment. Although this is necessary for the safeguarding of study staff and unlikely to exclude many individuals, this exclusion may miss those who are both highly vulnerable and potentially at the greatest risk of alcohol-related harm. Accurately quantifying alcohol consumption is challenging, especially among those who reported drinking home-brewed alcohol, where there is no standardised method of assessing the quality or percentage of alcohol.

In conclusion, the co-occurrence of alcohol use and *S. mansoni* infection presents a substantial public health challenge in rural Uganda. Our findings identify adult males and individuals engaged in fishing activity as high-risk groups, driven by a mix of social factors and concurrent with other lifestyle factors such as smoking. Future work is needed to investigate multi-sectoral public health interventions, for example integrating alcohol misuse screening into schistosomiasis control programmes, to address shared risk factors for alcohol use and *S. mansoni* infection and to assess their impact on health outcomes.

## DECLARATIONS

### Ethics Approval

Data collection and use were reviewed and approved by Oxford Tropical Research Ethics Committee (OxTREC 509-21), the Vector Control Division Research Ethics Committee of the Uganda Ministry of Health (VCDREC146), and the Uganda National Council for Science and Technology (UNCST HS1664ES).

## Acknowledgements

We thank the SchistoTrack group for their feedback, and Christin Puthur and Max Lang for their help with elements of the coding. We also give thanks to the field teams, including surveyors, technicians, nurses, sonographers, malacologists, and auxiliary workers. Special thanks go to the study participants within the 52 villages of SchistoTrack.

## Author contributions

Conceptualisation: BL, MEB, and GFC. Data curation: BL, BN, BT, JBO, NBK and GFC. Analysis: BL and MEB. Funding acquisition: BL and GFC. Investigation: BL, MEB. Methodology: BL, MEB, SL and GFC. Project administration: NBK and GFC. Resources: NBK and GFC. Supervision: SL and GFC. Writing – original draft: BL, MEB, and GFC. Writing – editing: BL, MEB, BN, BT, JBO, NBK, SL and GFC.

## Data availability

Individual participant data are not available due to the identifiable nature of the participant characteristics and ongoing nature of the cohort.

## Code availability

No new code was developed in the analysis of this study.

