## Supplementary material for "Co-occurrence of alcohol use and *Schistosoma mansoni* infection: prevalence, patterns, and risk factors in rural Uganda"

### Title

Prof. Goylette F. Chami

Big Data Institute

Old Road Campus

University of Oxford

Oxford, United Kingdom, OX3 7LF

**Supplementary methods**

Modifications to the WHO STEPS survey involved excluding questions on alcohol use that could not be recalled in this setting and revising the types of alcohol listed to ones which were appropriate to the location of the study, such as traditional spirits or homebrewed alcohol. The survey was validated during community engagement and training with local district staff in December 2021 before conducting the baseline of SchistoTrack. Questions on alcohol use were asked for participants aged 10 years and over. Participants were first asked whether they had ever consumed alcohol, coded as a categorical variable with options of yes, no, do not know. If yes, participants were then asked whether they had consumed alcohol in the past 12 months (yes, no, do not know). Individuals reporting alcohol use in the past year were further questioned on their drinking frequency (less than once per month, a few days per month, every week, every day, do not know), and what types of alcohol they consumed. Both variables were coded as categorical. Current alcohol users were defined as those reporting alcohol consumption within the past 12 months, otherwise considered current non-users. Current alcohol users were split into occasional alcohol users who consumed alcohol a few days per month or less and regular drinkers who consumed alcohol weekly or daily. Current non-users were split into life-time non-users who reported never consuming alcohol and former alcohol users who reported ever consuming alcohol, but not within the past year. Two participants who answered yes to ever consuming alcohol, but did not know whether they had consumed alcohol within the past 12 months were categorised as former alcohol users. Another four participants who reported consuming alcohol within the past 12 months, but did not know with what frequency were categorised as occasional alcohol users. This was based on the assumption that if they could not recall the frequency of their alcohol consumption, it was unlikely to be as frequent as weekly or daily.

### Supplementary figures

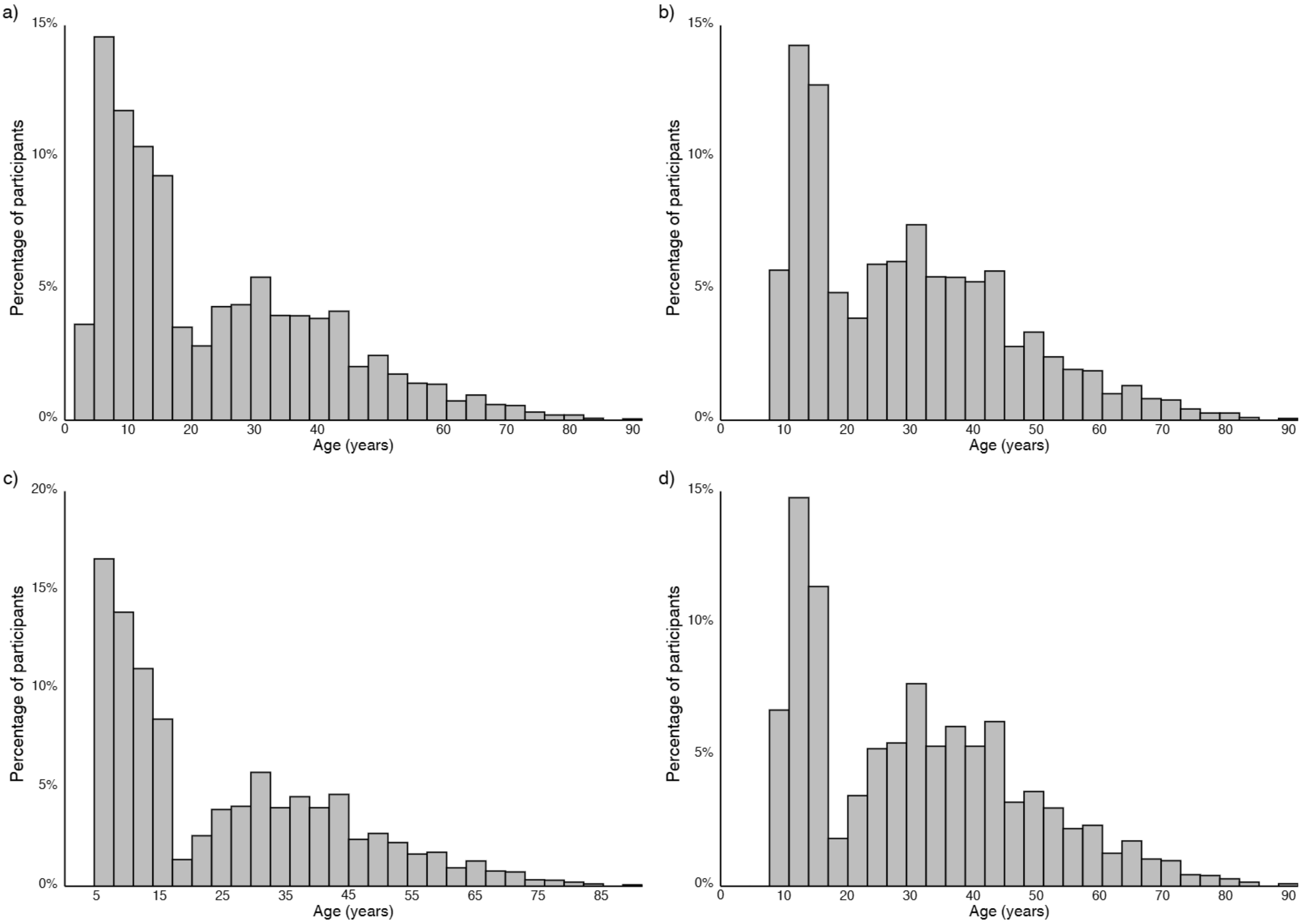


**Fig S1.** Age distributions for (a) all participants in the SchistoTrack cohort, (b) all participants with alcohol use data, (c) all participants with baseline *S. mansoni* infection data, and (d) all participants with both alcohol use data and baseline *S. mansoni* infection data.

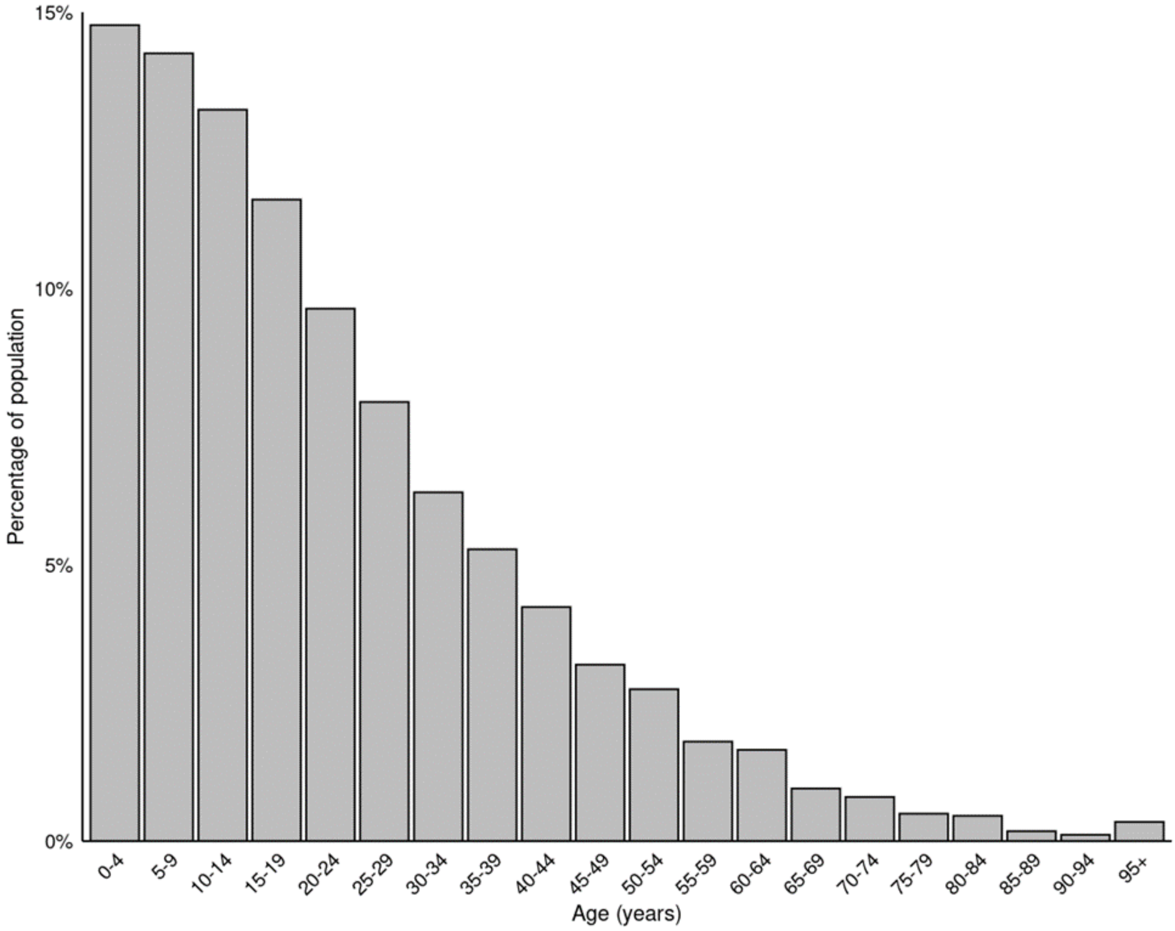


**Fig S2.** Distribution of ages according to the Ugandan National Housing and Population Census 2024, used for direct age standardisation.

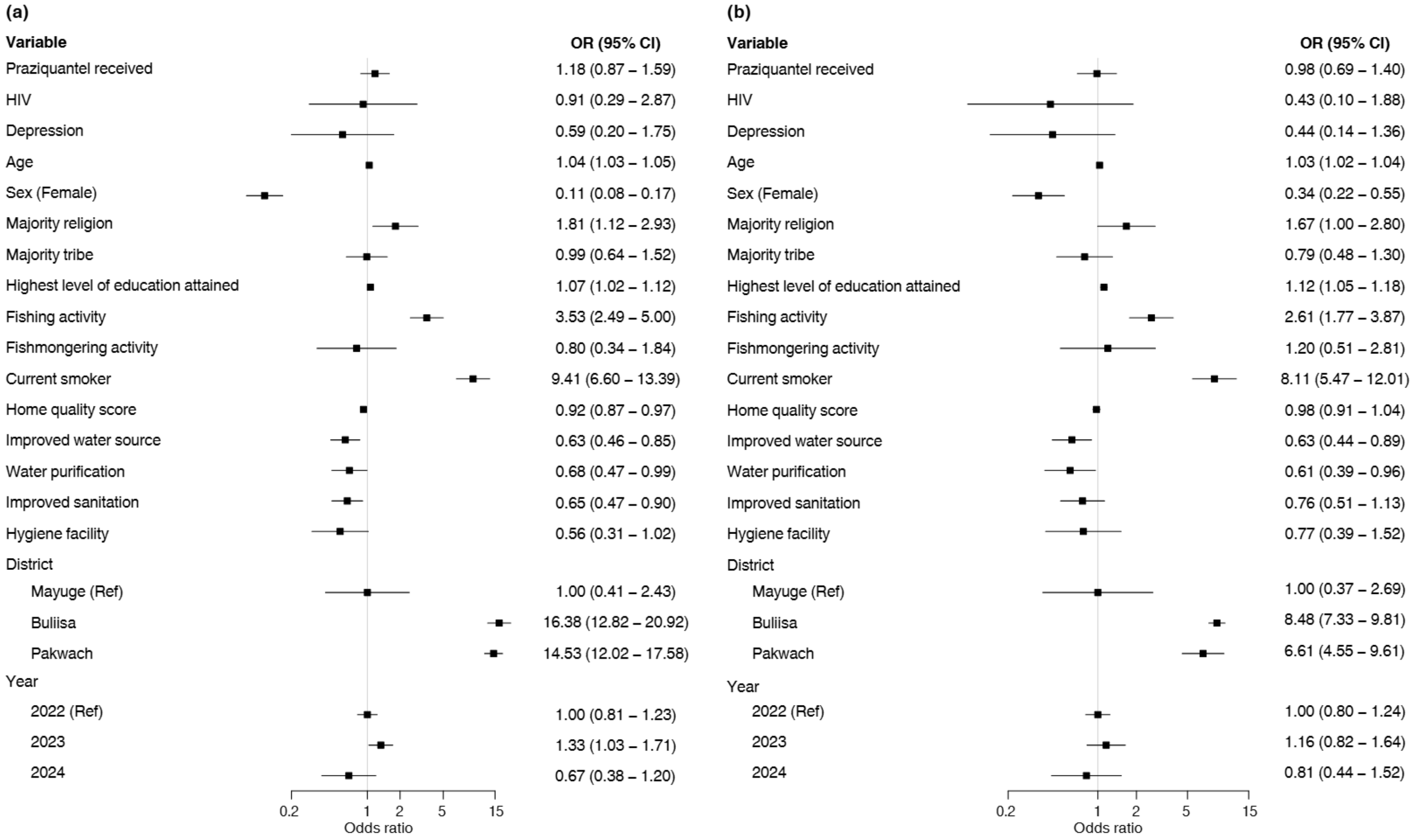


**Fig S3.** Models for regular alcohol use. Logistic regression models for regular alcohol use (214/3198), with floating absolute risks (FAR) shown for the district and year variables. (a) Minimally adjusted model, with each variable adjusted for age, sex, and district as appropriate. (b) Fully adjusted model including all covariates. Variance inflation factors (VIFs) were <5 for all variables. AUC for 10-fold cross-validation of the fully adjusted model was 0.895.

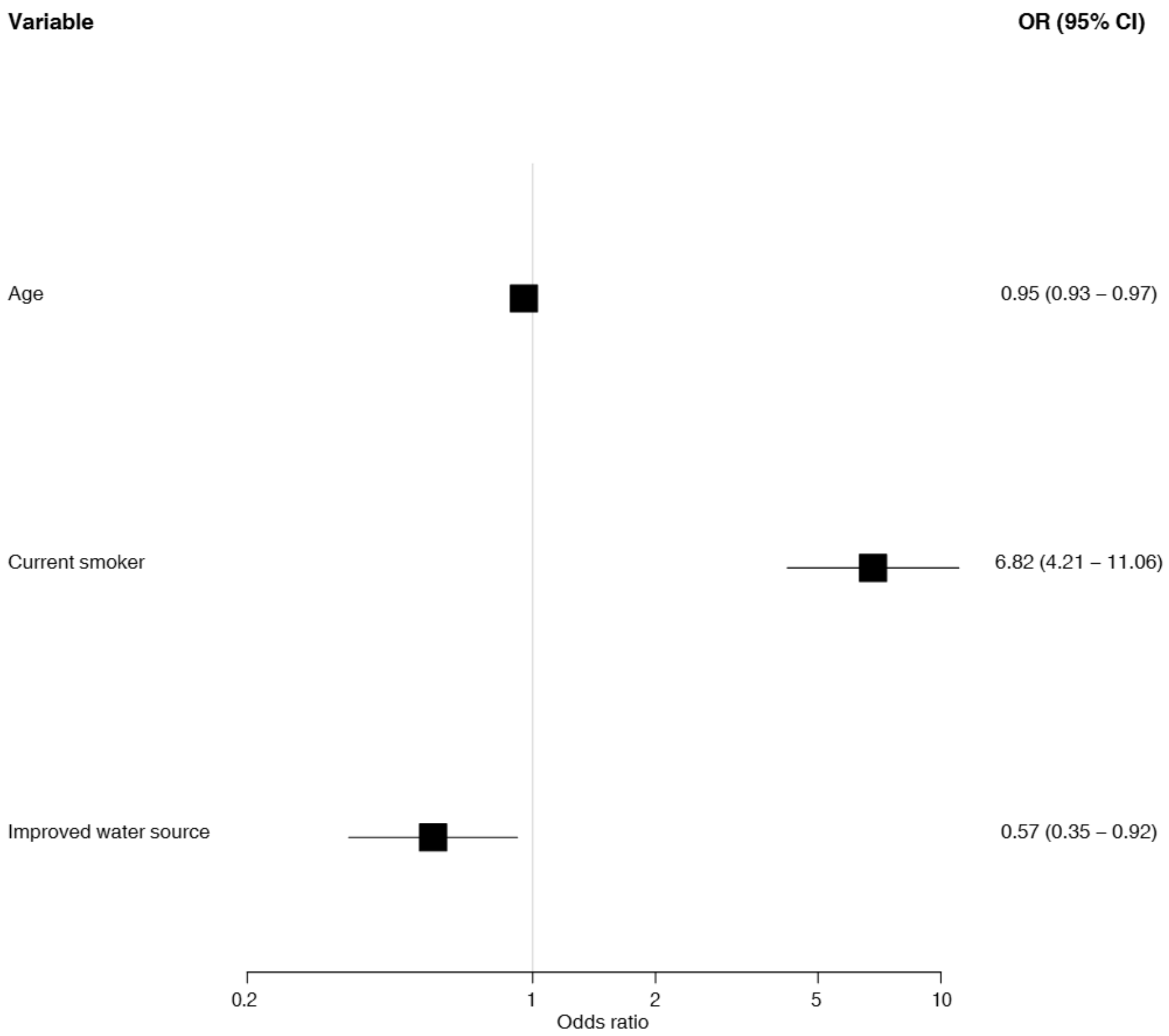


**Fig S4.** Logistic regression model for co-occurring regular alcohol use and *S. mansoni* infection among males aged ≥20 years (89/791). VIFs<5 for all variables. AUC for 5-fold cross-validation was 0.781.

**Table S5.** Table for infection intensities versus alcohol use group among males aged ≥20 years (N = 791).

| | *S. mansoni* infection category | | | | | | |
|---|---|---|---|---|---|---|---|
| **Alcohol consumption** | **No (n, %)** | **Low (n, %)** | **Mild (n, %)** | **High (n, %)** | **Total (n, %)** | **Chi-square (p value)** | **Spearman's rho** |
| **Alcohol non-user** | 288 (58.1) | 103 (21.0) | 60 (12.1) | 45 (8.8) | 496 (100) | 9.15 (0.17) | 0.093 (p = 0.0091) |
| **Occasional alcohol user** | 59 (50.0) | 28 (23.7) | 18 (15.3) | 13 (11.0) | 118 (100) | | |
| **Regular alcohol user** | 88 (49.7) | 36 (20.3) | 25 (14.1) | 28 (15.8) | 177 (100) | | |
| **Total** | 435 (55.0) | 167 (21.1) | 103 (13.0) | 86 (10.9) | 791 (100) | | |
